# Patterns of Crimean-Congo haemorrhagic fever virus seroprevalence in human and livestock populations in northern Tanzania

**DOI:** 10.1101/2023.08.31.23294720

**Authors:** Ellen C Hughes, William de Glanville, Tito Kibona, Blandina Theophil Mmbaga, Melinda K Rostal, Emanuel Swai, Sarah Cleaveland, Felix Lankester, Brian J Willett, Kathryn J Allan

**Author notes:** Corresponding author Ellen C Hughes, c/o MRC-University of Glasgow Centre for Virus Research, Glasgow, UK.

## Abstract

Results from a cross-sectional study of Crimean-Congo haemorrhagic fever virus (CCHFV) in northern Tanzania demonstrated high seroprevalence in humans and ruminant livestock with high levels of spatial heterogeneity. CCHFV may represent an unrecognised human health risk in this region and drivers of exposure need further investigation.

## Research Letter

Crimean-Congo haemorrhagic fever virus (CCHFV) is a tick-borne orthonairovirus with the potential to cause severe haemorrhagic fever in humans and for onward human-to-human transmission [1]. The virus is a World Health Organization priority pathogen for research and development [2]. A wide range of wild and domestic animals can be infected [3], but CCHFV does not typically cause clinical disease in non-human species [1]. In eastern Africa, since 2013 intermittent outbreaks of disease in humans have occurred in Uganda but the epidemiology remains poorly understood [4]. Northern Tanzania has been identified as an area likely to be at high risk of CCHF in humans [5] but no clinical cases have yet been reported in the country.

To investigate CCHFV exposure in northern Tanzania, we performed serological testing on human and ruminant livestock sera collected in 2016 using a multilevel sampling frame from 351 humans and 7456 livestock in linked households in Arusha and Manyara Regions (Figure 1) [6]. Sera were tested using a multi-species ELISA (ID Screen®, IDvet, Grabels, France) and seroprevalence estimated using the Survey package in R [7]. We assessed species-level differences in seroprevalence using a mixed-effects model with household and village as random effects. Patterns of spatial autocorrelation in village-level seroprevalence were investigated using Moran’s I statistic and correlation of village-level seroprevalence between species pairs was assessed using Pearson’s correlation coefficient (appendix).

**Figure 1:**
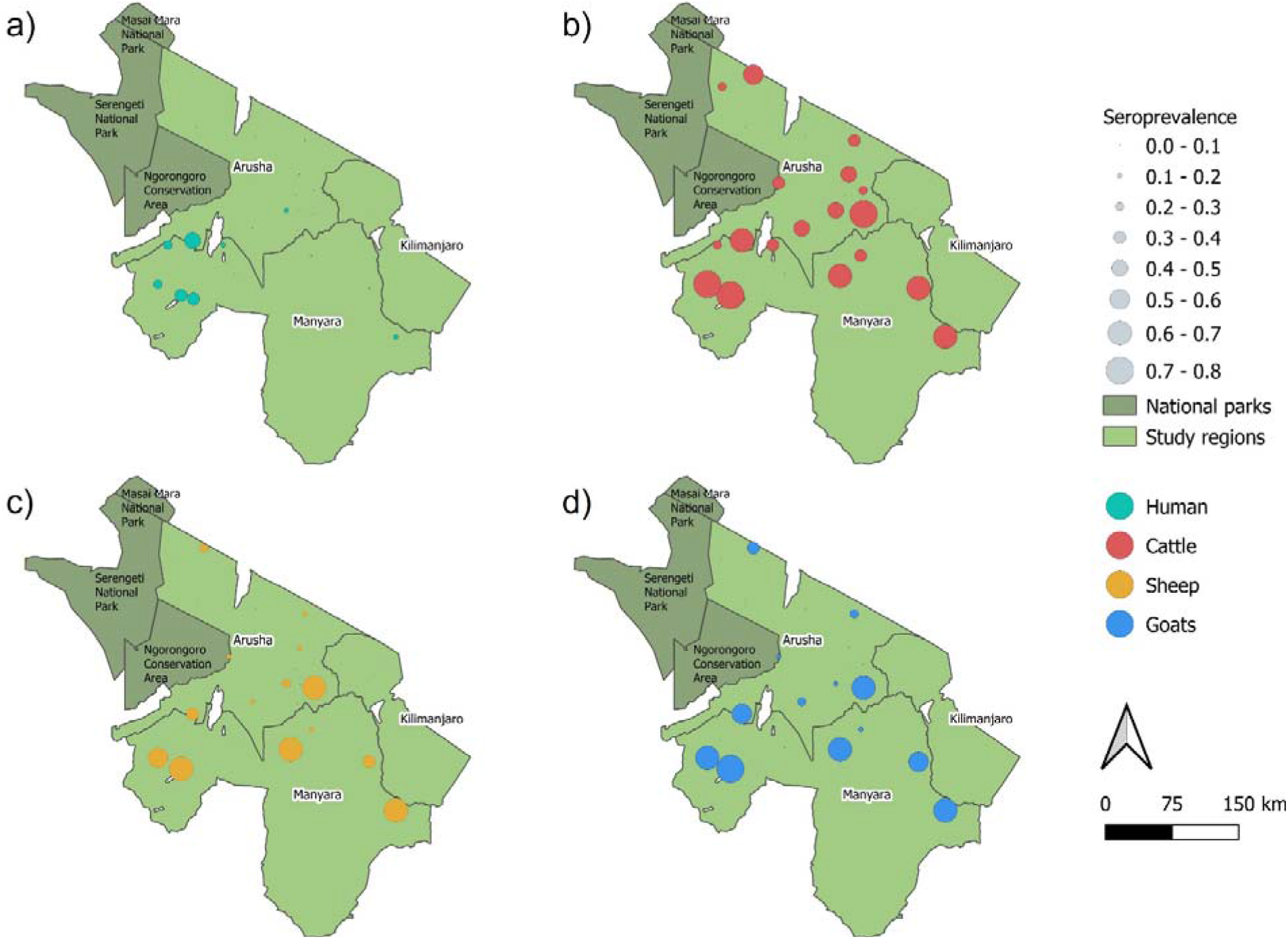
Seroprevalence of Crimean-Congo haemorrhagic fever virus for a) human, b) cattle, c) sheep and d) goats in villages in Arusha and Manyara regions, northern Tanzania

Overall seroprevalence was high in all livestock species (cattle: 49.6% (95% Confidence interval (CI) 40.0-59.2); goats: 33.8% (95% CI 21.7-47.5); sheep: 27.8% (95% CI 17.0-40.6) (Table 1, Figure 1), with sheep and goats having significantly lower odds of exposure than cattle (Sheep OR=0.32 (95% CI 0.27-0.37), p=<0.001; Goats OR=0.45 (95% CI 0.39-0.51), p=<0.001). Village-level seroprevalence ranged widely to a maximum of > 70% in each of the three livestock species (Table 1). While livestock seroprevalence can vary widely, these values were consistent with those reported elsewhere in East Africa [3]. The finding of a higher seroprevalence in cattle than in sheep and goats is also consistent with other settings in Africa [3] and may reflect differences in host feeding preferences of Hyalomma spp. ticks, considered to be important vectors of CCHFV [1]. However, further work is required to understand the relative contribution of different host species to viral maintenance, as well as their relationship to human infection risk.

**Table 1:** Seroprevalence of Crimean-Congo haemorrhagic fever virus in cattle, sheep, goats and humans in northern Tanzania. Number tested, overall seroprevalence, seroprevalence range by village and Moran’s I statistic for livestock and human serum samples collected in northern Tanzania in 2016 and tested for antibodies to Crimean-Congo haemorrhagic fever virus. Moran’s I statistic and associated p value are shown for the village level.

| Species | Tested (N) | Seroprevalence (95% CI) | Seroprevalence range by village (95% CI) | Moran's I statistic (p value) |
| --- | --- | --- | --- | --- |
|  |  |  |  | Village |
| Cattle | 3015 | 49.6 (40.0-59.2) | 5.3 (1.2-9.4) to 76.6 (70.3-82.8) | -0.09 (p=0.60) |
| Sheep | 2059 | 27.8 (17.0-40.6) | 0.0 (0-3.9) to 70.3 (55.5-85.0) | -0.09 (p=0.57) |
| Goats | 2382 | 33.8 (21.7-47.5) | 0.0 (0-3.9) to 70.3 (55.5-85.0) | -0.10 (p=0.61) |
| Human | 351 | 15.1 (8.5 – 23.8) | 0.0 (0.0-16.1) to 50 (30.7-69.2) | 0.43 (p=0.001) |

Overall human seroprevalence was 15.1% (95% CI 11.7-19.2) but village-level seroprevalence varied widely between study sites (Table 1). Seroprevalence was similar to that reported in health-care-seeking patients in Kenya in 2012 [10] but higher than the 1.2% values reported in community participants elsewhere in Tanzania [8]. However, given the substantial between-village variation observed (Table 1), interpretation of these regional comparisons is challenging.

Assessment of spatial autocorrelation via Moran’s I statistic (Table 1) showed no evidence of village-level spatial autocorrelation in livestock, suggesting that although context-specific drivers such as husbandry practices and local agro-ecology are likely important, drivers of exposure were not observable at a broader landscape level. In contrast, significant positive spatial autocorrelation was observed in the village-level human seroprevalence (Moran’s I statistic 0.43, p=<0.001). Additionally, species-pair correlations showed that village-level human and livestock seroprevalence were not correlated (rho=0.16 (p=0.51)), with high human seroprevalence seen in some low livestock prevalence locations and vice versa (Appendix). This heterogeneity, in combination with the differences in spatial distribution, suggest different possible drivers of exposure in livestock and human populations. For example, the infection prevalence of ticks in the peri-domestic environment may be a more important driver of infection for people than for livestock, which may be exposed across a wide range of environments when moving for grazing and water. However, as discrepancies in sample size may have exaggerated these differences, further linked investigation into human and livestock exposure and patterns of tick infection is required.

The high exposure levels to CCHFV in people implies that clinical CCHF is a potentially serious, underdiagnosed health risk in this population and suggests that CCHF should be included as a differential diagnosis for undifferentiated febrile illness in northern Tanzania. However, evidence of human seropositivity in the absence of clinical cases, even where health professionals are familiar with CCHF diagnosis, is common [9, 10]. The causes of disease emergence in such human populations are poorly understood and further research into regions such as northern Tanzania, where the virus is endemic but human disease has not been reported, is critical to understanding human disease risk.

Three key findings arise from this study: (a) CCHFV is circulating widely in both humans and livestock across northern Tanzania: (b) CCHFV seroprevalence shows high spatial heterogeneity and further investigations are needed to understand drivers of exposure; (c) high human seroprevalence demonstrates widespread exposure of people to the virus and suggests that CCHF should be included as a differential diagnosis for febrile illness in this region.

## Data Availability

All data produced in the present study are available upon reasonable request to the authors

## Conflict of interest

The authors declare no conflicts of interest.

## Notes

### Competing Interest Statement

The authors have declared no competing interest.

### Funding Statement

This research was supported by the Supporting Evidence Based Interventions project, University of Edinburgh (R83537) and the Zoonoses and Emerging Livestock Systems program (funded through BBSRC, DfID, ESRC, MRC, NERC and DSTL) (BB/L018926/1). EH was supported by the University of Glasgow, College of Medicine, Veterinary and Life Sciences Doctoral Training Programme.

### Author Declarations

Ethics committee of Kilimanjaro Christian Medical Centre (KCMC), Moshi, Tanzania (832) gave ethical approval for this work Ethics committee of National Institute of Medical Research (NIMR), Tanzania (2028) gave ethical approval for this work Ethics Committee of the University of Glasgow Medical, Veterinary and Life Sciences (MVLS) (200140152) gave ethical approval for this work. Permission to carry out the study in Tanzania was provided by the Tanzania Commission for Science and Technology (2014-244-ER-2005-141). Written informed consent or assent for sample collection and questionnaire administration was collected from all participants.

