## appendix for "Patterns of Crimean-Congo haemorrhagic fever virus seroprevalence in human and livestock populations in northern Tanzania"

### Study design

Samples were collected as part of a cross-sectional sero-survey undertaken in 2016 to investigate several zoonotic pathogens in livestock and humans across a range of agricultural systems [1, 2]. Livestock samples were collected using a multilevel sampling approach with 3015 cattle, 2382 goats and 2059 sheep sampled from 417 households in 20 villages randomly selected using generalized random tessellation stratified sampling to ensure spatial balance [2, 3]. Human serum samples (n=351) were collected from a random selection of 113 of these households across 17 villages, as well as four additional village sites without linked livestock samples. As GPS co-ordinates were not recorded for these sites, they do not appear on the maps (Figure 1).

### Laboratory testing

Samples from all species were heat-treated at 56 °C for two hours in Tanzania before shipping to the UK (TARP(S)2016/49) for analysis at the MRC-University of Glasgow Centre for Virus Research using a commercially produced, species-independent, double antigen sandwich ELISA (IDvet, Grabels, France). Manufacturer-reported sensitivity and specificity values for the ID Screen® CCHF Double Antigen Multi-species (IDvet, Grabels, France) are shown in Table A1.

***Table A1 Manufacturer-reported sensitivity and specificity values for the ID Screen® CCHF Double Antigen Multi-species (IDvet, Grabels, France)***

| | **Specificity** | | **Sensitivity** | |
|---|---|---|---|---|
| **Species** | **Specificity (%** | **95% confidence intervals** | **Sensitivity (%)** | **95% confidence intervals** |

| | | | | |
|---|---|---|---|---|
| **Cattle** | **100** (n = 402) | 99.1 - 100 % | **97.9** (n = 95) | 92.6 – 99.4 % |
| **Sheep** | **100** (n = 402) | 99.1 - 100 % | **99.0** (n = 102) | 94.7 – 99.8 % |
| **Goats** | **100** (n = 402) | 99.1 - 100 % | **100** (n = 74) | 95.1 – 100 % |
| **Humans** | **100** (n = 257) | 98.5 - 100 % | *Not available* | - |

### Statistical analysis

Seroprevalence was calculated for each species both at a population and village-level. Overall seroprevalence and 95% confidence intervals were calculated using the *Survey* package in R, using village and household as cluster identifiers (village = primary sampling unit, household = secondary sampling unit) [4]. Village-level seroprevalence was calculated as the number of positives/total number sampled in each village with binomial confidence intervals.

Village-level prevalence for all species was plotted on maps of the study area. Maps were created in QGIS (version 3.16.0 [5]). All statistical analyses were performed in R statistical environment, version 3.6.1 [6]. Mixed-effects logistic regression models were implemented using the *lme4* package [7]. Moran's I was calculated using the *spdep* package in R [8]. A Moran's I statistic of 1 is equivalent to perfect spatial clustering, while a value of -1 represents perfect dispersal. Statistical significance was set at $p \leq 0.05$.

### Ethics

The study protocols, questionnaires, and consent documents were approved by the Kilimanjaro Christian Medical Centre (KCMC) (832) and National Institute of Medical Research (NIMR) (2028) ethics committees, and University of Glasgow Medical, Veterinary and Life Sciences (MVLS) Ethics Committee

(200140152). Permission to carry out the study in Tanzania was provided by the Tanzania Commission for Science and Technology (2014-244-ER-2005-141). Written informed consent or assent for sample collection and questionnaire administration was collected from all participants. Samples were imported into the UK under license TARP(S)2016/49. Permission to publish was granted by the Director of Veterinary Services, Tanzania(Act No 17 of 2003).

### Results

*Table A2 Odds of exposure to CCHFV in sheep and goats compared to cattle. Odds ratio (OD), 95% confidence intervals and p values from an all-species mixed effect logistic regression model with species as a fixed effect, and village and household as random effects are shown.*

| Variable | Level | Odds Ratio (OR) | OR 95% confidence intervals | | p value |
|---|---|---|---|---|---|
| | (Intercept) | 0.75 | 0.41 | 1.36 | 0.341 |
| Species | Cattle | Reference | | | |
| | Goat | 0.45 | 0.39 | 0.51 | <0.001 |
| | Sheep | 0.32 | 0.27 | 0.37 | <0.001 |

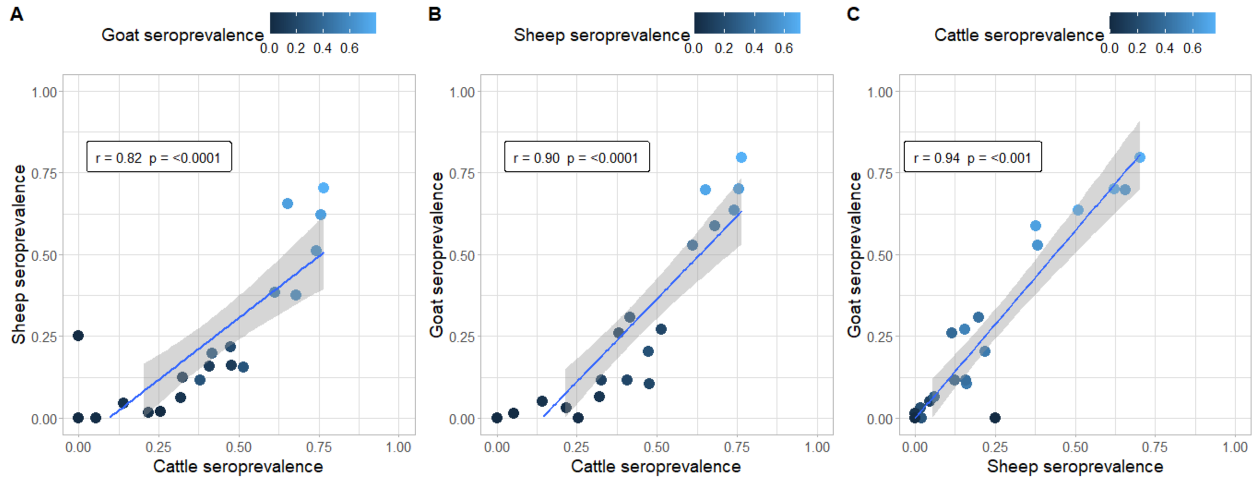


*Figure A1 Correlation between village-level seroprevalence by species-pairs. a) cattle and sheep, b) cattle and goats, c) sheep and goats. Points are coloured according to the village-level log odds of the third species.*

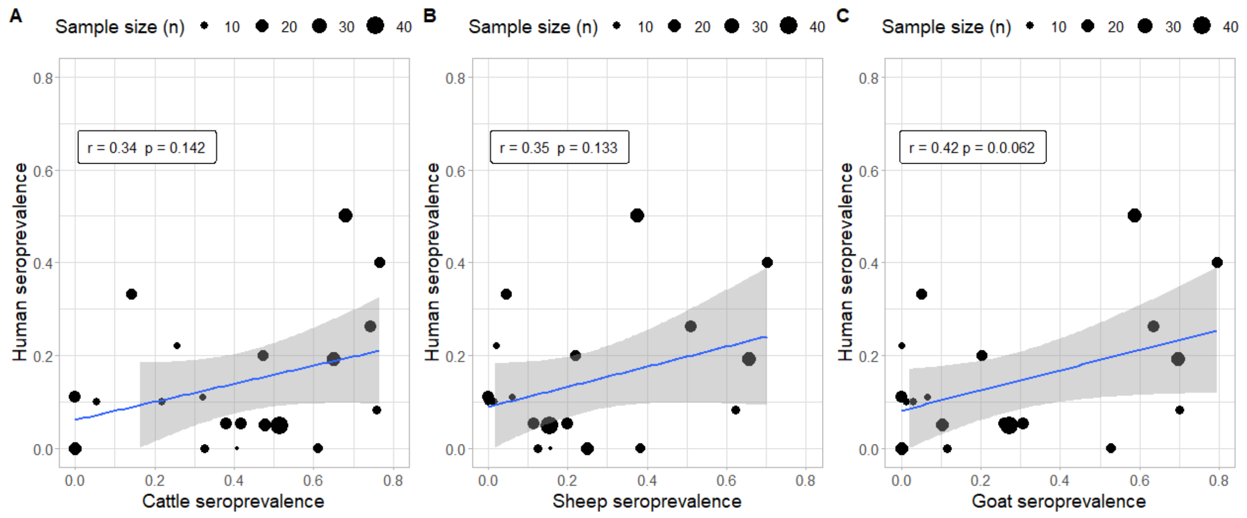


Figure A2 Correlation between village-level human and livestock seroprevalence. a) human and cattle, b) human and sheep, c) human and goats. Points are sized according to the village-level sample size.

### Acknowledgements

We would like to thank the livestock-keepers who participated in this study, as well as village, ward, district and regional authorities. We are grateful to Kunda Mnzava, Tauta Maapi, Rigobert Tarimo, Fadhili Mshana, Zanuni Kweka, Euphrasia Mariki, Ephrasia Hugho, Nelson Amani, Victor Mosha, and Elizabeth Kasagama for their contribution to field and/or laboratory work. This research was supported by the Supporting Evidence Based Interventions project, University of Edinburgh (R83537) and the Zoonoses and Emerging Livestock Systems program (funded through BBSRC, DfID, ESRC, MRC, NERC and DSTL) (BB/L018926/1). EH was supported by the University of Glasgow, College of Medicine, Veterinary and Life Sciences Doctoral Training Programme.
